# Using Feedback on Symptomatic Infections to Contain the Coronavirus Epidemic: Insight from a SPIR Model

**DOI:** 10.1101/2020.04.14.20065698

**Authors:** Michael Nikolaou

**Affiliations:** Chemical & Biomolecular Engineering Department, University of Houston, Houston, TX 77204-4004

**Keywords:** COVID-19, Coronavirus, Flattening the Curve, Social Distancing, Population Dynamics, Control

## Abstract

A study is presented on the use of real-time information about symptomatic infectious individuals to adjust restrictions of human contacts at two basic levels, the stricter being on the symptomatic infectious group. Explicit analytical formulas as well as numerical results are presented to rapidly elucidate what-if questions on averting resurgence of the coronavirus epidemic after the first wave wanes. Implementation of related ideas would rely on a mix of several factors, including personal initiative and sophisticated technology for monitoring and testing. For robust decision making on the subject, detailed multidisciplinary studies remain indispensable.

## 1. Introduction

In the absence of a vaccine, restrictions on social distancing have been used widely in recent efforts to avert an inordinately large number of fatalities from potentially uncontrolled spread of COVID-19.^1-3^ Underlying these efforts are fundamental constraints emerging from the dynamics of infectious disease spreading, as captured in the most elementary form by the standard SIR model:^4-7^ The coronavirus basic reproductive ratio, *R*_0_, naturally between 2 and 3 (Figure 1)^8^ should be brought close to 1 (or below) by social distancing measures,^8,9^ with correspondingly flattened curves of percent-infectious over time.^10-12^

**Figure 1.**
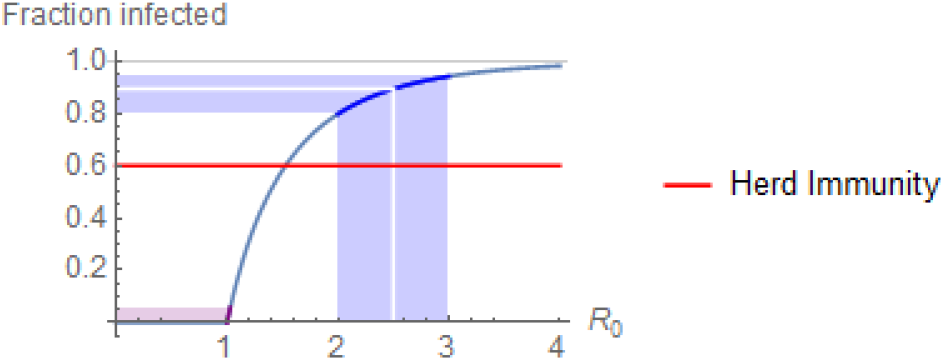
*Total fraction of a population infected, r*(∞) *– thus rendered immune – by the end of an epidemic, as a function of the basic reproductive ratio R*_0_, *according to the solution* 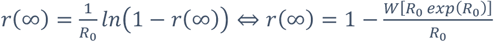 *of the SIR model (similar to eqn.(9)). For R*_0_ < 1 *the virus does not spread and the epidemic is contained. The infectious fraction reduces over time when the population fraction susceptible to infection is below* 1 − 1/*R*_0_ ≈ 60% *for the coronavirus*.

With the end of the first wave of the epidemic projected in a few weeks, questions arise about removal of social distancing measures. As desirable as the “return to normal” is, fundamental constraints remain that prevent complete removal of restriction measures until a vaccine is available for safe use.^13^ Indeed, the total fraction of infected – therefore eventually immune – by the end of the first wave will be at single percentage points,^2^ far below the 1 − 1/*R*_0_ ≈ 60% required to robustly avert resurgence of the epidemic.^7^ Therefore, a number of strategies are considered, such as alternating imposition and removal of measures over time,^1^ stratification by age, pre-existing conditions, location, or other risk-factors,^1,14^ mass screening, contact tracing, testing of all individuals entering the country, and quarantine of people who test positive.^15^

The purpose of this paper is to analyze the dynamics of simply placing restrictions on people infected by the virus *after* they have shown *symptoms* of the disease. No testing is required during an individual’s pre-symptomatic period, although such testing would certainly be beneficial, as that pre-symptomatic individual would be infective immediately upon infection.^16^ Restrictions may be self-imposed and assisted by telecommunications technology, a practice already implemented in Asian countries and under consideration in the US.^17^

In the rest of the paper, the basic analytical results are derived and illustrated by numerical simulations.

## 2. Containing virus spread by monitoring symptoms

The SPIR model structure (Figure 2) comprises population fractions susceptible to the virus, *s*; pre-symptomatic infectious, *p*; symptomatic infectious, *i*; and removed from the infectious pool, *r*, by recovery or death. A distinction between the SPIR model structure and other four-compartment structures, such as the classic SEIR,^4-7,18^ is that individuals with coronavirus infection entering the P group can infect before symptoms appear. Therefore, it is practically formidable to monitor those infecting during the pre-symptomatic period, as this would require inordinately massive testing or widespread tracking of contacts to guide selective testing. On the other hand, monitoring individuals with symptoms is more reasonable, as symptoms are fairly characteristic of the infection and testing for confirmation of the infection can be highly targeted.^19^ This type of interaction between the S, I, and P compartments is captured in the model structure shown in Figure 2. This structure indicates dynamics by separately adjustable feedback from the I group, in addition to feedback from the P group typically considered.

**Figure 2.**
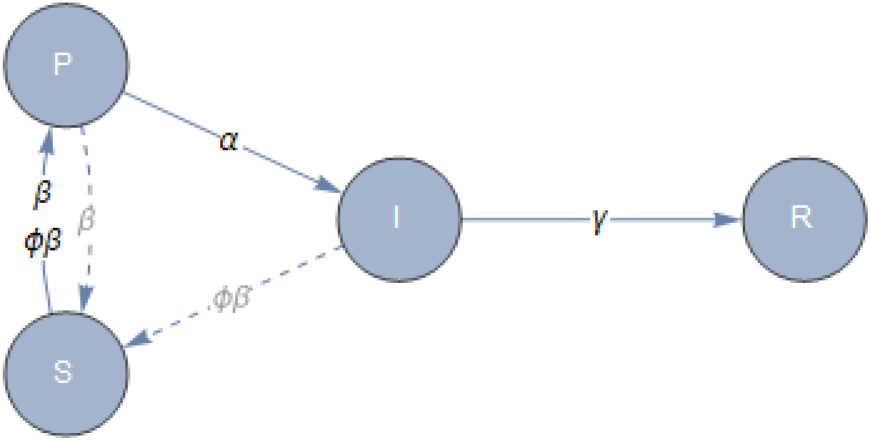
*The SPIR model structure, with the pre-symptomatic infectious group*, P, *and the symptomatic infectious group*, I, *infecting the susceptible group*, S, *at different rates, due to different distancing measures*.

Consequently, the following equations capture the dynamics of a fixed-size population with measures restricting transmission of the virus between susceptible and symptomatic infectious individuals:

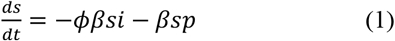

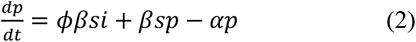

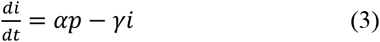

where β is the infection spread rate; 0 ≤ ϕ < 1 is a factor producing a reduced spread rate, ϕβ, between susceptible and symptomatic infectious; *α, γ* are the removal rates from the pre-symptomatic and symptomatic infectious groups, respectively; and 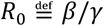 is the basic reproductive ratio. The fourth fraction, *r* = 1 − *s* − *p* − *i*, refers to individuals removed from the infectious group, by either recovery or death. Clearly, then,

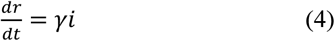

Based on eqns. (1)-(4) it can be easily shown (*Appendix A*) that herd immunity is achieved when the susceptible fraction, *s*, is less than

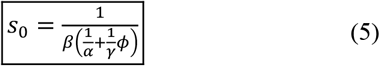

with obvious increase of *s*_0_ towards *α*/ *β* as *ϕ* → 0.

If no additional measures are taken to mitigate virus transmission from symptomatic infectious to susceptible, i.e. ϕ = 1, the resulting *s*_0_ becomes

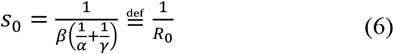

where 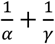 refers to the average time from becoming infected to removal from the infected group. Eqn. (6) implies that to keep *s*_0_ = 1 − *x* ≈ 1, uniform restrictions on virus transmission must achieve

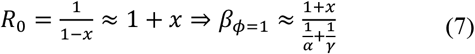

However, with additional measures taken to mitigate virus transmission from symptomatic infectious to susceptible, i.e. 0 ≤ *ϕ* = 1, eqn. (6) implies that the resulting *ϕ* becomes

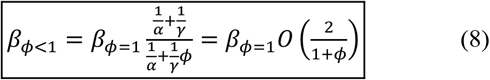

assuming *α* and *γ* are of the same order of magnitude.^20^ In the extreme, if unrealistic, case of perfect detection and full quarantine for the symptomatic infectious (*ϕ* = 0), restrictions on the rest of the population would be about half the size of equivalent uniform restrictions on the entire population (*ϕ* = 1).

It should be noted that eqns. (1)-(4) suggest that even perfect restriction of virus transmission from the symptomatic infectious alone cannot yield adequate results, as the pre-symptomatic infectious would be enough to infect the population at numbers not lower enough than those without any restrictions. Indeed, it can be shown (*Appendix B*) that the total fraction of the population infected by the end of the epidemic, *r*(∞), satisfies the equation

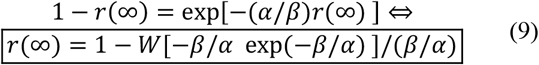

(where *W* is the *Lambert function*^21,22^)* with

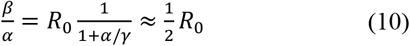

for *α* ≈ *γ*, which in turn yields a total fraction of infected by the end of the epidemic around 60% (Figure 1). Therefore *coordinated reduction of both ϕ and ϕ (restrictions both for the symptomatic infectious and for the rest of the population, respectively) would be required for desirable results*.

Given a growing body of data that can be used to estimate the effect of corresponding policies on the resulting β,^9,23^ the above simple analysis can help gauge restriction measures at the early conceptual level of making structural decisions.

## 3. Simulations

The values 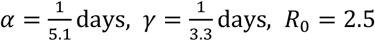 are used in all simulations.^20^ The well known basic case of doing nothing to contain the virus is shown in Figure 3, for reference.

**Figure 3.**
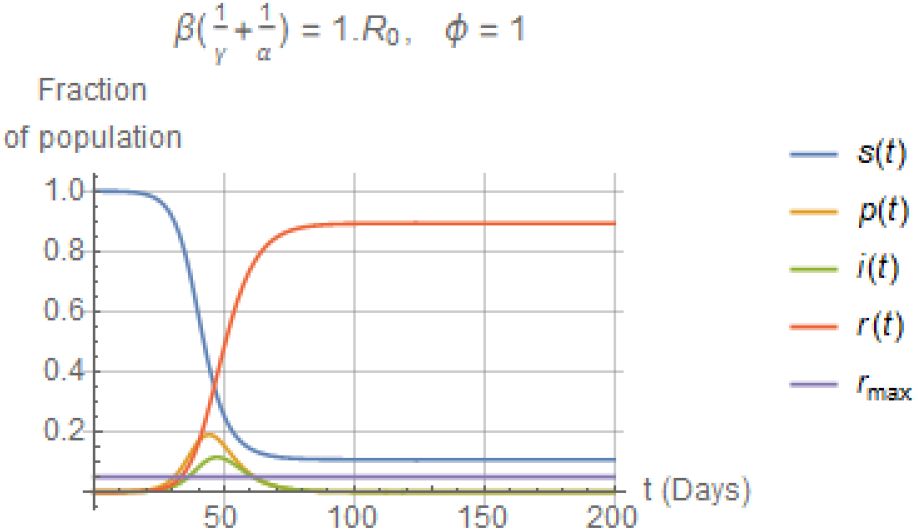
*Basic case of progress of the coronavirus epidemic in the absence of any restrictions on the population. The cumulative fraction of infected in the epidemic, equal to r*(∞), *would be about 0*.*9, in agreement with Figure 1 for R*_0_ = 2.5.

In the next case, the symptomatic infectious part of the population is placed on quarantine, *ϕ* = 0 (Figure 4).

**Figure 4.**
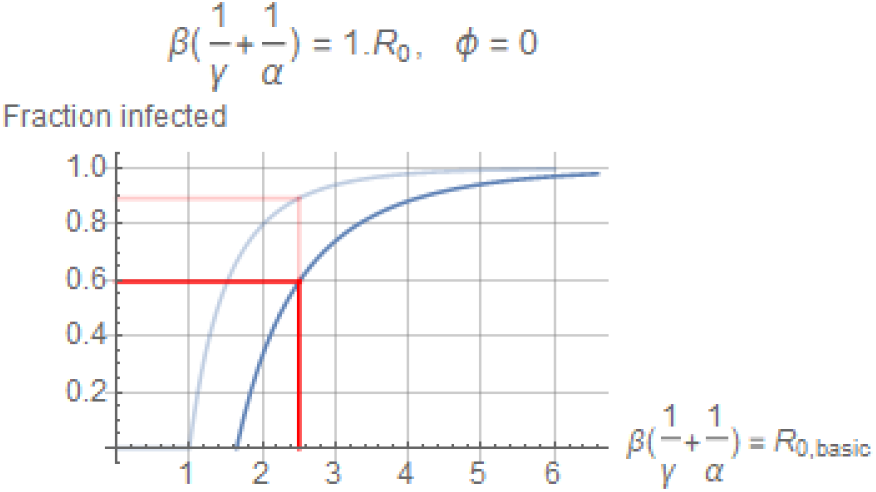
*Placing the symptomatic infectious on quarantine without placing any restrictions on the rest of the population would yield r*(∞) = 1 − *W*[−(*β*/*α*) *exp*(−*β*/*α*)]/(*β*/*α*) = 0.6. *The curve shown in Figure 1 is also included for comparison*.

Finally, placing restrictions on both the symptomatic infectious and the rest (0.6*R*_0_) with tighter restrictions on the former (*ϕ* = 0.3) yields quite improved results (Figure 6).

**Figure 5.**
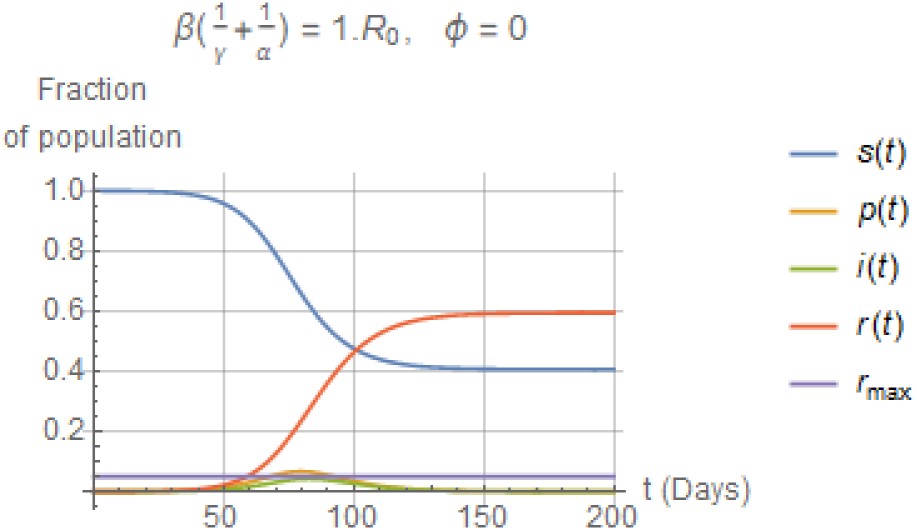
*Placing the symptomatic infectious on quarantine would yield r*(∞) = 1 − *W*[−(*β*/*α*) *eexp*(−*β*/*α*)]/(*β*/*α*) = 0.6 *as suggested by Figure 4*.

**Figure 6.**
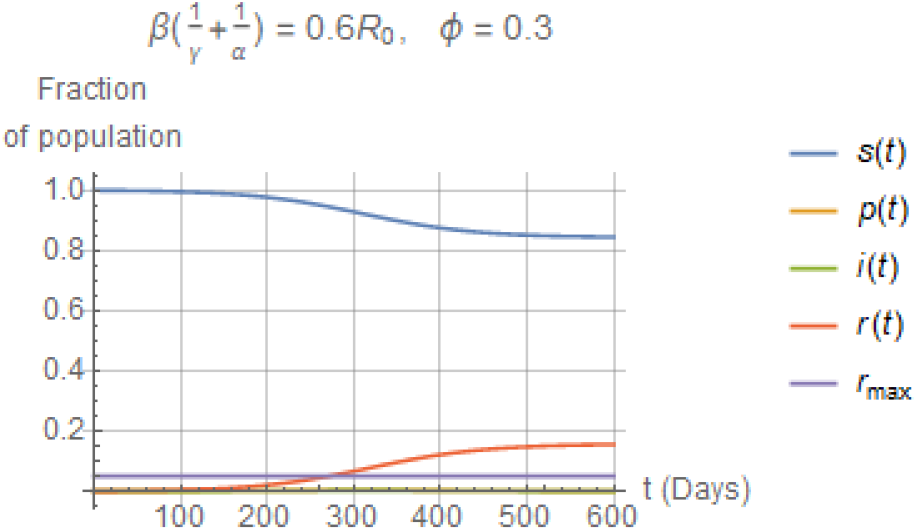
Placing restrictions on both the symptomatic infectious and the rest of the population with tighter restrictions on the former yields significantly improved results.

The final value *r*(∞) suggested by eqns. (9) and (10), shown in Figure 4, is confirmed by the profile of *r*(*t*) shown in Figure 5. The improvement over Figure 3 is inadequate.

Note the quite inferior outcomes, were tighter restrictions not placed on symptomatic infectious (*ϕ* = 1, Figure 7).

**Figure 7.**
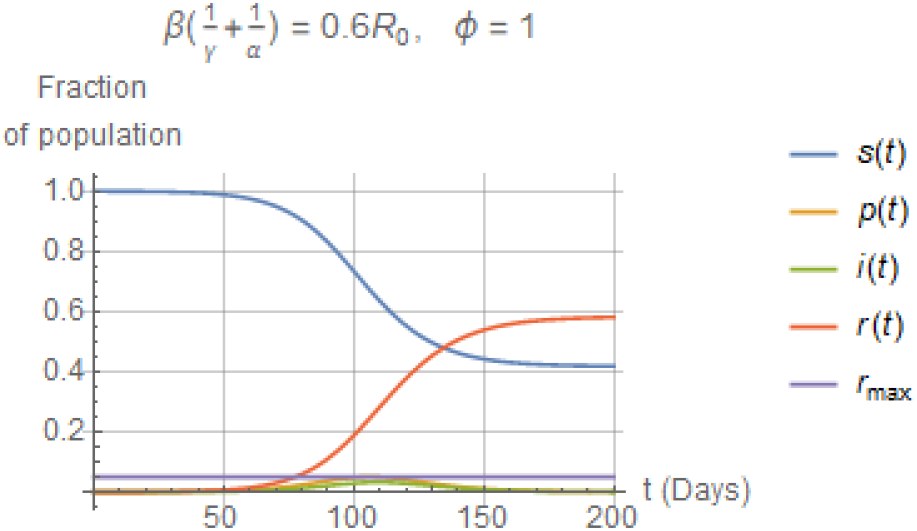
Not placing restrictions on the symptomatic infectious while leaving the rest of the population at the same restriction level yields far inferior outcomes.

## 4. Discussion

A study was presented on the use of real-time information about symptomatic infectious individuals to adjust restrictions of human contacts in areas affected by coronavirus infections. To the extent that measures can be taken to effectively implement such restrictions, they could help effective containment of the epidemic, particularly if combined with ideas tailoring measures to account for risk stratification. As already pointed out, successful implementation would hinge on a mix of several factors, including personal initiative and sophisticated technology for monitoring and testing. The simple formulas and graphs presented here, based on widely available background, will hopefully help provide guidelines for rapidly addressing what-if questions in a transparent way. For robust decision making detailed multidisciplinary studies remain indispensable.^24^

## Data Availability

Not applicable

## 5. Acknowledgements

All computations were done in *Mathematica*, available at the University of Houston. Sharing of teaching material about the SIR model on Github by Prof. Jeff Kantor of Notre Dame is also gratefully acknowledged.

## Appendix A.

*Proof of eqn. (5)*

Eqns. (2) and (3) imply

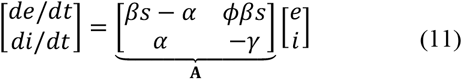

The eigenvalues of the matrix **A** are in the left half-plane iff and

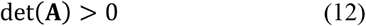

and

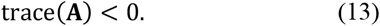

Eqn. (12) implies

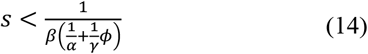

Eqn. (13) implies

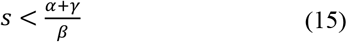

which is trivially guaranteed by eqn. (14), leading to eqn. (5).

## Appendix B.

*Proof of eqn. (9)*

Standard analysis of eqns. (1) and (2) for *ϕ* = 0 proceeds as follows:

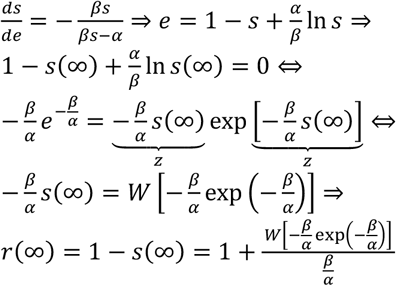

## Footnotes

* *Interestingly, the analytical solution for r*(∞) *in terms of the Lambert function W*(*z*), *eqn.(9), pointed out as early as 1996*,^21^ *may have escaped the attention of most literature in this field*.^7^

